# An Updated Evidence Assessment of the Genetic Causes of Dilated Cardiomyopathy

**DOI:** 10.64898/2026.03.09.26347990

**Authors:** Elizabeth Jordan, Phoenix Grover, Patricia Parker, Jason Cowan, Babken Asatryan, Tomohiko Ai, Akos Berthold, Lucas Bronicki, Emily Brown, Rudy Celeghin, Mat Edwards, Judith Fan, Cindy James, Renee Johnson, Daniel P. Judge, Sean Jurgens, Najim Lahrouchi, Tom Lumbers, Francesco Mazzarotto, Argelia Medeiros Domingo, Brittney Murray, Stacey Peters, Kalliopi Pilichou, Alexandros Protonotarios, Karin van Spaendonck-Zwarts, Petros Syrris, Jessica Wang, Roddy Walsh, James Ware, Ray E. Hershberger

**Author notes:** Co-Corresponding Authors: Elizabeth Jordan MS, CGC, The Ohio State University Wexner Medical Center, Biomedical Research Tower Room 306, 460 West 12^th^ Avenue, Columbus, Ohio USA 43210. or Ray E. Hershberger, MD, The Ohio State University Wexner Medical Center, Biomedical Research Tower Room 304, 460 West 12^th^ Avenue, Columbus, OH 43210 USA.

## Abstract

**Background:** Evidence of the diverse genetic architecture of dilated cardiomyopathy (DCM) continues to emerge and requires reassessment of the clinical relevance of implicated disease genes. Building on the 2019-2020 Clinical Genome Resource (ClinGen) evaluation, the DCM Gene Curation Expert Panel (GCEP) reconvened in 2024-2025 to conduct a reassessment of genes in DCM.

**Methods:** The ClinGen semi-quantitative clinical validity classification framework was applied with specifications to DCM to classify genes into categories based upon strength of published evidence for a DCM phenotype. Previously curated genes were reassessed and newly reported gene-disease-mode of inheritance (MOI) relationships, termed “curations,” were evaluated.

**Results:** Sixty-eight genes were evaluated, inclusive of 72 unique gene-disease-MOI relationships across 51 previously evaluated and 17 newly assessed genes. Thirty-five curations were classified as high evidence (16 Definitive, 10 Strong, 9 Moderate), increasing by 16 from the prior assessment. Nine newly assessed genes were classified as high evidence, including *BAG5, FLII, LMOD2, MYLK3, MYZAP, NRAP, PPA2, PPP1R13L,* and *RPL3L*. Twelve genes (11 newly appraised) were rated as high evidence with an autosomal recessive (AR) MOI. Five re-evaluated genes from 2019-2020 had clinically significant changes in classification. Except for *JPH2*, for which curation was modified to separate autosomal dominant (-AD) and -AR MOI curations, clinically significant changes involved upgrades from low to high evidence categories (*PLEKHM2, PRDM16, TBX20, TNNI3K*), demonstrating the robustness of the ClinGen gene curation process over time. An additional 29 gene-disease-MOI curations were classified as Limited, including six newly evaluated genes and one new MOI for a previously evaluated gene, *MYBPC3-*AR; four were classified as No Known Disease Relationship, and four remained Disputed. Four previously evaluated genes were curated for both AD and AR MOIs, including *JPH2* (AD-Strong, AR-Limited), *LDB3* (AD-Limited, AR-Strong), *MYBPC3* (AD-Limited, AR-Limited), and *TNNI3* (AD- and AR- Strong).

**Conclusions:** With substantial new evidence, the genetic architecture of DCM has rapidly expanded. This updated assessment of genes reported in DCM yielded 35 high evidence curations, an increase from 19 only five years ago. The results of this evidence-based evaluation process informs clinical interpretation of genetic information in the care of DCM patients and families.

**CLINICAL PERSPECTIVE:** *What’s new?:*

- The Clinical Genome Resource (ClinGen) Dilated Cardiomyopathy (DCM) Gene Curation Expert Panel reconvened to update the evidence for genes in DCM using the ClinGen clinical validity framework.
- A total of 35 genes were classified into clinically actionable, high evidence categories of Definitive, Strong, or Moderate evidence, an increase of 16 from the 2019-2020 curation.
- Of the 16 newly classified high evidence curations, 11 (69%) had an autosomal recessive mode of inheritance and were observed primarily in pediatric DCM.

*What are the clinical implications?:*

- This update has substantially expanded the complex and diverse genetic architecture of DCM spanning 18 gene ontologies (8 new) identified in both adult and pediatric patients.
- The 19 genes classified as high evidence in the 2019-2020 curations were adopted by the clinical genetics community as the key genes for clinical genetic testing and care for DCM patients and families. The 35 high evidence curations from the current assessment are recommended to be used as an updated list for clinical genetics care for DCM.
- DCM gene curation will require ongoing reassessments due to the continuing expansion of high-quality research data.

## INTRODUCTION

A great deal of new evidence has emerged since the Clinical Genome Resource (ClinGen)^1, 2^ Dilated Cardiomyopathy (DCM) Gene Curation Expert Panel (GCEP) first curated genes for strength of evidence supporting a causative relationship with DCM.^3^ In just five years, an unexpectedly large number of genes not previously curated have had evidence published suggesting a relationship with DCM. To date, published efforts curating the strength of evidence for gene-disease relationships for cardiomyopathies has followed the classic clinical phenotypes of dilated, hypertrophic, and arrhythmogenic right ventricular cardiomyopathies (DCM, HCM, ARVC).^4^ HCM has an estimated prevalence of 1/500.^5^ Most of the rare variant genetic basis of HCM has been identified in genes that encode proteins of the sarcomere, with *MYBPC3* and *MYH7* contributing to 85-90% of identifiable genetic cause, which reaches >95% when considering contributions from *TNNI3*, *TNNT2* and *TPM1.*^6^ Other less common genetic causes of left ventricular hypertrophy have been recently curated and defined.^7^ Similarly, most of the identifiable genetic basis of classical ARVC, with an estimated prevalence of 1/5000,^8^ stems from rare variants impacting the desmosome, a key structure for cardiac myocyte adhesion and electrical function. Three genes, *PKP2*, *DSP*, and *DSG2*, contribute most of the genetic causes of ARVC with *DSC2*, *JUP*, and *TMEM43* less common contributors.^9^

DCM is the most common cardiomyopathy with a prevalence estimated at 1/250^10^ to 1/220.^11^ DCM contributes an outsized degree to heart failure with reduced ejection fraction, a major public health problem,^12^ which underscores the significance of understanding the genetic basis of DCM. In the 2019-2020 ClinGen evaluation, 19 genes from 10 ontologies, biological and molecular functional groups,^13^ were classified as high evidence demonstrating the diversity of DCM genetic etiology.^3^ Aside from rare variants in *TTN* and *LMNA*, contributing 10-15% and 4-5% of identified genetic cause of DCM, respectively, the contributions of the numerous other genes to DCM cause are much less. Nevertheless, mechanistic insights derived from an expanded DCM gene ontology may fuel novel insights for diagnostic, treatment, and preventive strategies for DCM and heart failure.

Per the ClinGen re-curation timeline procedures,^1, 2^ the DCM GCEP was reconvened with international and broad clinical and genetics expert representation to again undertake a rigorous, equipoise- and data-driven approach to evaluate previously curated and newly identified candidate genes implicated in DCM. Here we report the updated evidence-based appraisal of genes in their monogenic relationship with DCM.

## METHODS

The data that support the findings of this study are publicly available and published on the ClinGen website (https://clinicalgenome.org/affiliation/40035/). ClinGen curations are updated periodically. To find the most current information visit clinicalgenome.org. As a systematic analysis of clinical and experimental data, no formal statistical testing was performed. This research does not involve human subjects and therefore Institutional Review Board approval was not required.

The methods used for this work built upon previously published approaches.^3, 14, 15^ Established panel members re-assembled as the DCM GCEP, with new members (P.G., P.P., J.C., A.B., S.J., K.S.) added to enhance expertise areas where prior members were no longer able to participate. The GCEP implemented the ClinGen gene-disease validity classification standards^15^ using the current Standard Operating Procedures with specifications to DCM (Supplemental Material). Biweekly teleconference meetings to curate the data were held from May 3, 2024 through May 30, 2025.

To generate the initial gene list, a structured literature search and gene-disease reference resource inquiry was performed as of January 1, 2024, using the same approach implemented from the initial assessment.^3^ The initial list identified 406 genes for evaluation, an increase from the 267 genes from the prior effort. Each gene was evaluated to select those with evidence suggesting a plausible relationship to monogenic, non-syndromic DCM. Additional details on panel membership, operational implementation, and development and triaging of the gene list can be found in the Supplemental Material.

### Phenotype Definition

The DCM phenotype was defined by systolic dysfunction, conventionally noted as a left ventricular (LV) ejection fraction of <50% accompanied by LV enlargement, after other usual clinically detectable causes of cardiomyopathy were excluded, as previously defined and applied for this purpose.^3^ In cases where LV systolic dysfunction without LV enlargement was observed, points at a reduced grade were considered if, in the opinion of the clinical experts on the panel, the assessment of the complete clinical data were consistent with DCM. Publications used for scoring of clinical genetic evidence were required to specify how the DCM phenotype was defined and that other clinically detectable causes were excluded. In the absence of such specifications, the data was either not scored or scores were substantially reduced.

Some genes included in this report were curated for disease names identified by disease names (e.g., syndromic) extending beyond DCM alone and/or were primarily curated by other ClinGen expert panels and labeled with MONDO^16,17^ identifiers accordingly. . These genes are also recognized in this report because DCM evidence contributed a significant number of points to the final determinations: *ACTN2* (MONDO:0700349, *ACTN2-*related cardiac and skeletal myopathy), *PLN* (MONDO:00005091, intrinsic cardiomyopathy), *PPP1R13L* (MONDO:0957795, arrhythmogenic cardiomyopathy with variable ectodermal abnormalities), *DSP* (MONDO:0011581, arrhythmogenic cardiomyopathy with wooly hair and keratoderma), and *NKX2-5* (MONDO:0800441, *NKX2.5-*related congenital, conduction, and myopathic heart disease).

### Gene Curation and Evidence Scoring Process

Each unique gene-disease-mode of inheritance (MOI) combination individually evaluated is referred to herein as a “curation.” For each curation, the ClinGen framework sums scores for published clinical genetic and experimental laboratory evidence. “Genetic evidence” included clinical data, encompassing variant, segregation, and case-control analyses, when available. The default scoring and adjustment approaches for variant-level evidence was specified for DCM and depended on molecular consequence and MOI (autosomal dominant (AD) and autosomal recessive (AR)) under curation (Supplemental Material). For example, the GCEP elected to not score individuals or pedigrees with >1 possibly relevant variant in any putative DCM gene. In addition, homozygous variants arising in a confirmed or suspected consanguineous family were systematically reduced in points due to potential confounding factors that could be associated with excess homozygosity. Experimental evidence was assessed by category (e.g., expression data, functional alterations, model systems, and rescue).

The ClinGen clinical validity classifications include *Definitive and Strong* (12-18 points), *Moderate* (7-11 points), *Limited* (1-6 points), *No Known Disease Relationship* (NKDR), *Disputed*, and *Refuted*.^1, 2^ The maximum number of genetic evidence points that could be given was 12, and the maximum number of experimental evidence points was 6, for a highest total possible score of 18 points. “Definitive” is defined as a curation with a Strong evidence score with ≥2 independent reports over 3 years and no contradictory evidence. If the point-based classification was not considered to reflect the collective assessment of the panel’s clinical and scientific interpretation of the data or if the final point value fell between categories (between 6-7 points Limited versus Moderate; 11-12 points Moderate versus Strong/Definitive), the classification was further discussed on a convened call with panel members. When applicable, the final classification was modified to a category reflective of consensus of the panel’s expert adjudication of the collective evidence. All evidence evaluated and rationale for determinations were entered into the ClinGen Gene Curation Interface. Data entered can be viewed on the public ClinGen website.^18^ Additional details on the evidence evaluation, specifications, and scoring procedures are provided in the Supplemental Material and have been previously published.^3, 15^

New curations and updated curations of genes previously classified as Moderate or Strong were presented to the full panel on conference calls to establish consensus for an approved clinical validity classification. For genes previously evaluated as low evidence (Limited or NKDR) and not proposed to change classification, two independent experts conducted expedited reviews with feedback and approval collected by email. If no change in final classification was made upon agreement by the curators and experts, the final score and written evidence summary was circulated to the full panel for approval. Alternatively, for any proposed change in classification or if there was discordance between the recommendations of the experts upon offline review, the evidence was presented and final classification discussed with the panel on a convened call to determine a final classification.

When applying the American College of Medical Genetics and Genomics (ACMG) and Association for Molecular Pathology (AMP) variant interpretation standards,^19^ prevailing guidance holds that variants in genes with high evidence classifications (Definitive/Strong or Moderate) may achieve clinically actionable pathogenic or likely pathogenic (P/LP) classifications, whereas variants identified in low evidence genes (Limited, NKDR) are generally classified as variants of uncertain significance (VUS).^20^ Accordingly, care was taken at expert review, understanding that gene assignments of Limited versus Definitive, Strong, or Moderate have clinically relevant implications.

## RESULTS

Sixty-eight genes were identified for assessment: 17 new and 51 re-appraised. In addition, four of the 51 previously evaluated genes had new MOIs assessed. When counting each curation individually, a total of 72 curations were conducted. Thirty-five were classified as high evidence (16 Definitive, 10 Strong, 9 Moderate), 29 Limited, four NKDR, and four remained Disputed (Table 1). Ten of the re-appraised curations changed classification, five of which were clinically significant, including four upgrades and one downgraded due to new evidence warranting separate AD and AR curations (Table 2). Curations classified into high evidence categories spanned 18 gene ontologies (Figure 1), an increase from 10 from the prior effort.^3^ The genetic and experimental evidence points contributing to each classification achieving Limited, Moderate, or Strong/Definitive are shown in Figure 2.

**Figure 1.**
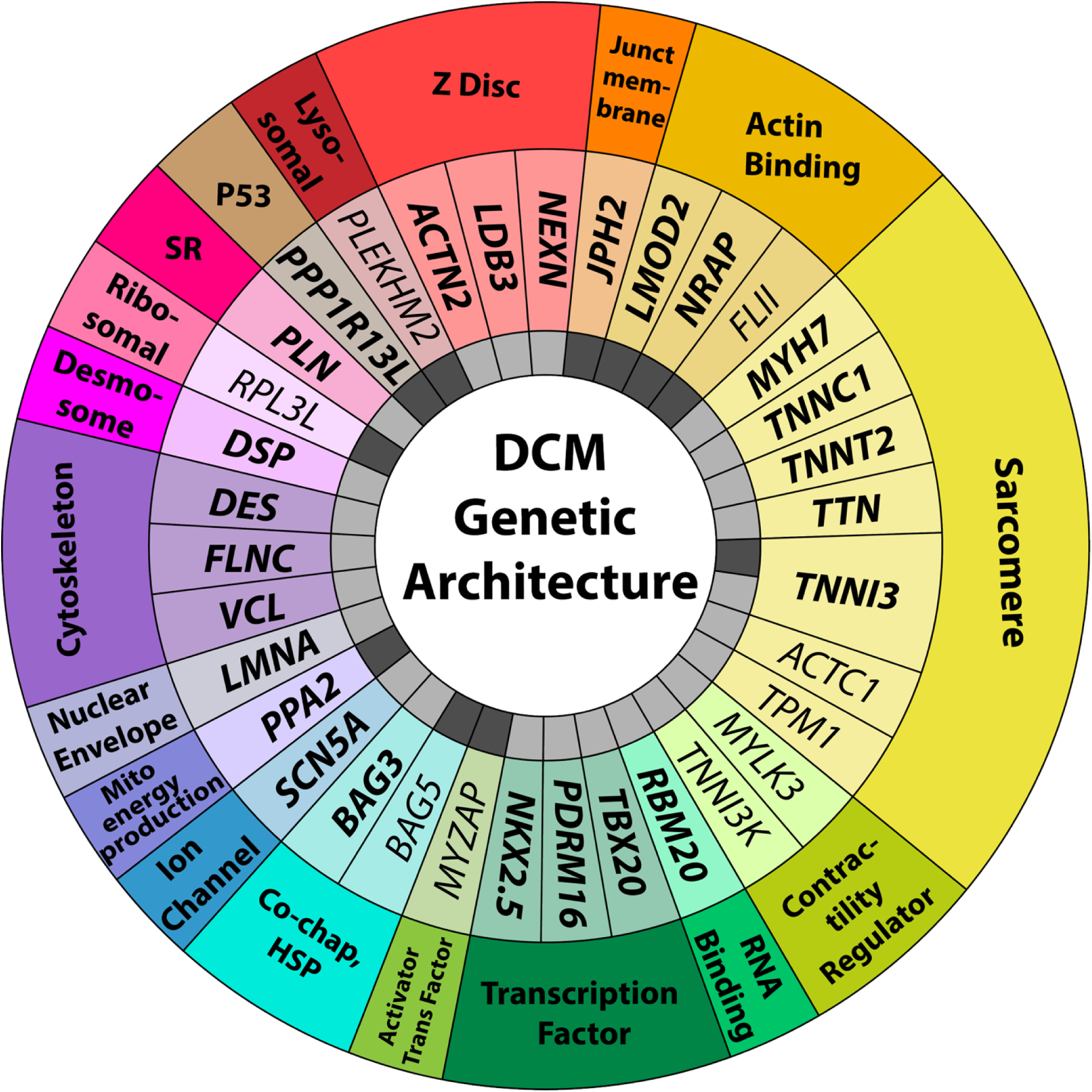
The genetic architecture of dilated cardiomyopathy. The innermost circle indicates the mode of inheritance associated with each high evidence gene as a result of the curation for DCM: Light Gray = Autosomal Dominant and Dark Gray = Autosomal Recessive. *TNNI3* has both light and dark gray inner ring cell colors due to having two unique Strong classification curations for dominant and recessive inheritance, respectively. The center circle depicts all gene names with high evidence classifications of Definitive, Strong, or Moderate. Bolded gene names are Definitive or Strong evidence. The genes are grouped by gene ontology, defined herein as similar biological, molecular, and/or protein functional groups, as noted in the outermost circle, with abbreviations defined as follows: Co-Chap, HSP = Co-Chaperone, Heat Shock Protein; Junct Membrane = Junctional Membrane; Mito Energy Production = Mitochondrial Energy Production; P53= P53 inhibitor; SR= Sarcoplasmic Reticulum.

**Figure 2.**
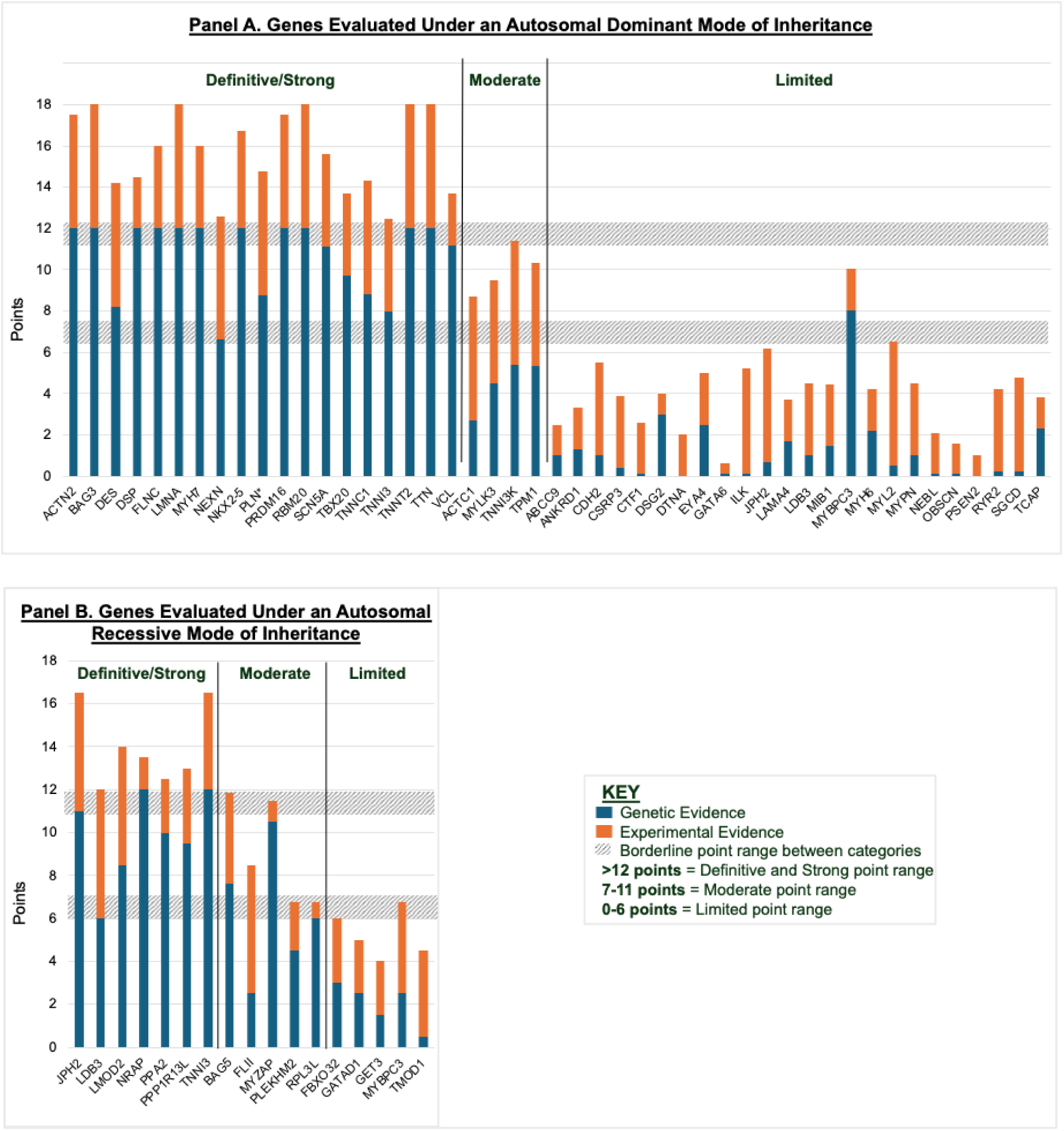
Quantitative contributions of genetic and experimental evidence to the clinical validity classifications of the genes classified as Limited, Moderate, Strong or Definitive. Blue is clinical genetic evidence contributions and orange is experimental evidence contributions to the total quantitative score for each gene-disease-mode of inheritance curation. The gray bars demonstrate the point thresholds distinguishing Limited and Moderate evidence score (lower bar, with values of 6-7 points falling in between these categories) and Moderate and Strong/Definitive evidence score (upper bar, with values of 11-12 points falling in between these categories). If point values exceed these lines as defined, it was the opinion of the expert panel that the quantitative scores did not reflect the overall qualitative assessment of the evidence and the final classification category was modified despite the accumulation of points for the final score. Genes noted with an asterisk were primarily scored by another GCEP within the ClinGen Cardiovascular Domain working group.

**Table 1.** Quantitative Scores and Final Classifications of Evaluated Gene-Disease-Mode of Inheritance Curations.

| Gene | Protein | MOI | Genetic Evidence | Experimental Evidence | Total Score | Curation Type | Classification |
| --- | --- | --- | --- | --- | --- | --- | --- |
| *ACTN2* * | Alpha-actinin-2 | AD | 12 | 5.5 | 17.5 | Recuration | Definitive |
| *BAG3* | BCL2-associated anthanogene 3 | AD | 12 | 6 | 18 | Recuration | Definitive |
| *DES* | Desmin | AD | 8.2 | 6 | 14.2 | Recuration | Definitive |
| *DSP* * | Desmoplakin | AD | 12 | 2.5 | 14.5 | Recuration | Definitive |
| *FLNC* | Filamin C | AD | 12 | 4 | 16 | Recuration | Definitive |
| *LMNA* | Lamin A | AD | 12 | 6 | 18 | Recuration | Definitive |
| *LMOD2* | Leiomodin 2 | AR | 8.5 | 5.5 | 15 | New | Definitive |
| *MYH7* | Myosin heavy chain 7 | AD | 12 | 4 | 16 | Recuration | Definitive |
| *NKX2-5* * | NK2 homeobox 5 | AD | 12 | 4.75 | 16.75 | Recuration | Definitive |
| *PLN* * | Phospholamban | AD | 8.75 | 6 | 14.75 | Recuration | Definitive |
| *PPP1R13L* | Protein phosphatase 1 regulatory subunit 13 like | AR | 9.5 | 3.5 | 13 | New | Definitive |
| *RBM20* | RNA-binding motif protein 20 | AD | 12 | 6 | 18 | Recuration | Definitive |
| *SCN5A* | Sodium voltage-gated channel, alpha subunit 5 | AD | 11.1 | 4.5 | 15.6 | Recuration | Definitive |
| *TNNC1* | Troponin C | AD | 8.8 | 5.5 | 14.3 | Recuration | Definitive |
| *TNNT2* | Troponin T2 | AD | 12 | 6 | 18 | Recuration | Definitive |
| *TTN* | Titin | AD | 12 | 6 | 18 | Recuration | Definitive |
| *JPH2* | Junctophilin 2 | AR | 11 | 5.5 | 16.5 | New | Strong |
| *LDB3* | LIM domain-binding 3 | AR | 6 | 6 | 12 | New | Strong |
| *NEXN* | Nexilin F-actin-binding protein | AD | 6.6 | 6 | 12.6 | Recuration | Strong |
| *NRAP* | Nebulin related anchoring protein | AR | 12 | 1.5 | 13.5 | New | Strong |
| *PPA2* | Inorganic pyrophosphatase 2 | AR | 10 | 2.5 | 12.5 | New | Strong |
| *PRDM16* | PR domain-containing protein, family M, member 2 | AD | 12 | 5.5 | 17.5 | Recuration | Strong |
| *TBX20* | T-box transcription factor 2- | AD | 9.7 | 4 | 13.7 | Recuration | Strong |
| *TNNI3* | Troponin I3 | AD | 7.95 | 4.5 | 12.45 | Recuration | Strong |
| *TNNI3* | Troponin I3 | AR | 12 | 4.5 | 16.5 | New | Strong |
| *VCL* | Vinculin | AD | 11.2 | 2.5 | 13.7 | Recuration | Strong |
| *ACTC1* | Actin alpha cardiac muscle 1 | AD | 2.7 | 6 | 8.7 | Recuration | Moderate |
| *BAG5* | BCL-associated anthanogene 5 | AR | 7.6 | 4.25 | 11.85 | New | Moderate |
| *FLII* | FLII actin remodeling protein | AR | 2.5 | 6 | 8.5 | New | Moderate |
| *MYLK3* | Myosin light chain kinase 3 | AD | 4.5 | 5 | 9.5 | New | Moderate |
| *MYZAP* | Myocardial zonula adherens protein | AR | 10.5 | 1 | 11.5 | New | Moderate |
| *PLEKHM2* | Pleckstrin homology domain-containing protein 16 | AR | 4.5 | 2.25 | 6.75 | Recuration | Moderate |
| *RPL3L* | Ribosomal protein L3 like | AR | 6 | 0.75 | 6.75 | New | Moderate |
| *TNNI3K* | TNNI3-interacting kinase | AD | 5.4 | 6 | 11.4 | Recuration | Moderate |
| *TPM1* | Tropomyosin | AD | 5.35 | 5 | 10.35 | Recuration | Moderate |
| *ABCC9* | ATP-binding cassette, subfamily C, member 9 | AD | 1 | 1.5 | 2.5 | Recuration | Limited |
| *ANKRD1* | Ankryn repeat domain-containing protein 1 | AD | 1.3 | 2 | 3.3 | Recuration | Limited |
| *CDH2* | Cadherin 2 | AD | 1 | 4.5 | 5.5 | New | Limited |
| *CSRP3* | Cysteine and glycine-rich protein 3 | AD | 0.4 | 3.5 | 3.9 | Recuration | Limited |
| *CTF1* | Cardiotrophin 1 | AD | 0.1 | 2.5 | 2.6 | Recuration | Limited |
| *DSG2* | Desmoglein 2 | AD | 3 | 1 | 4 | Recuration | Limited |
| *DTNA* | Dystrobrevin alpha | AD | 0 | 2 | 2 | Recuration | Limited |
| *EYA4* | EYA transcriptional coactivator and phosphatase 4 | AD | 2.5 | 2.5 | 5 | Recuration | Limited |
| *FBXO32* | F-box protein 32 | AR | 3 | 3 | 6 | New | Limited |
| *GATA6* | GATA binding protein 6 | AD | 0.1 | 0.5 | 0.6 | New | Limited |
| *GATAD1* | Gata zinc finger domain-containing protein 1 | AR | 2.5 | 2.5 | 5 | Recuration | Limited |
| *GET3* | Guided entry of tail-anchored proteins factor 3, ATPase | AR | 1.5 | 2.5 | 4 | New | Limited |
| *ILK* | Integrin linked kinase | AD | 0.1 | 5.1 | 5.2 | Recuration | Limited |
| *JPH2* | Junctophilin 2 | AD | 0.7 | 5.5 | 6.2 | Recuration | Limited |
| *LAMA4* | Laminin alpha 4 | AD | 1.7 | 2 | 3.7 | Recuration | Limited |
| *LDB3* | LIM domain-binding 3 | AD | 1 | 3.5 | 4.5 | Recuration | Limited |
| *MIB1* | Mindbomb E3 ubiquitin protein<br>ligase 1 | AD | 1.45 | 3 | 4.45 | Recuration | Limited |
| *MYBPC3* | Myosin binding protein C | AR | 2.5 | 4.25 | 6.75 | New | Limited |
| *MYBPC3* | Myosin binding protein C | AD | 8.05 | 2 | 10.05 | Recuration | Limited |
| *MYH6* | Myosin heavy chain 6 | AD | 2.2 | 2 | 4.2 | Recuration | Limited |
| *MYL2* | Myosin light chain 2 | AD | 0.5 | 6 | 6.5 | Recuration | Limited |
| *MYPN* | Myopalladin | AD | 1 | 3.5 | 4.5 | Recuration | Limited |
| *NEBL* | Nebulette | AD | 0.1 | 2 | 2.1 | Recuration | Limited |
| *OBSCN* | Obscurin | AD | 0.1 | 1.5 | 1.6 | Recuration | Limited |
| *PSEN2* | Presenilin 2 | AD | 0 | 1 | 1 | Recuration | Limited |
| *RYR2* | Ryanodine receptor 2 | AD | 0.2 | 4 | 4.2 | New | Limited |
| *SGCD* | Sarcoglycan delta | AD | 0.25 | 4.5 | 4.75 | Recuration | Limited |
| *TCAP* | Titin-cap | AD | 2.3 | 1.5 | 3.8 | Recuration | Limited |
| *TMOD1* | Tropomodulin 1 | AR | 0.5 | 4 | 4.5 | New | Limited |
| *LRRC10* | Leucine rich repeat containing 10 | AR | 0 | 4.25 | 4.25 | Recuration | NKDR |
| *NPPA* | Natriuretic peptide precursor A | AR | 0 | 0.75 | 0.75 | Recuration | NKDR |
| *RTKN2* | Rhotekin 2 | AR | 0 | 0 | 0 | New | NKDR |
| *VEZF1* | Vascular endothelial zinc finger 1 | AD | 0 | 1 | 1 | New | NKDR |
| *MYL3* | Myosin light chain 3 | AD | 0.1 | 0.5 | 0.6 | Recuration | Disputed |
| *PDLIM3* | PDZ and LIM domain 3 | AD | 0 | 0 | 0 | Recuration | Disputed |
| *PKP2* | Plakophilin 2 | AD | 0.2 | 0.5 | 0.7 | Recuration | Disputed |
| *PSEN1* | Presenilin 1 | AD | 0 | 0.5 | 0.5 | Recuration | Disputed |

MOI=Mode of inheritance; AD=autosomal dominant; AR= autosomal recessive; Recuration=a curation that was previously evaluated/published and evidence was re-evaluated in this project; New=a curation that was newly evaluated not previously published for DCM that was assessed in this project; NKDR=no known disease relationship; \*Other GCEPs within the ClinGen Cardiovascular Disease Working Group performed and/or contributed to the final scores and classifications of these curations.

**Table 2.**
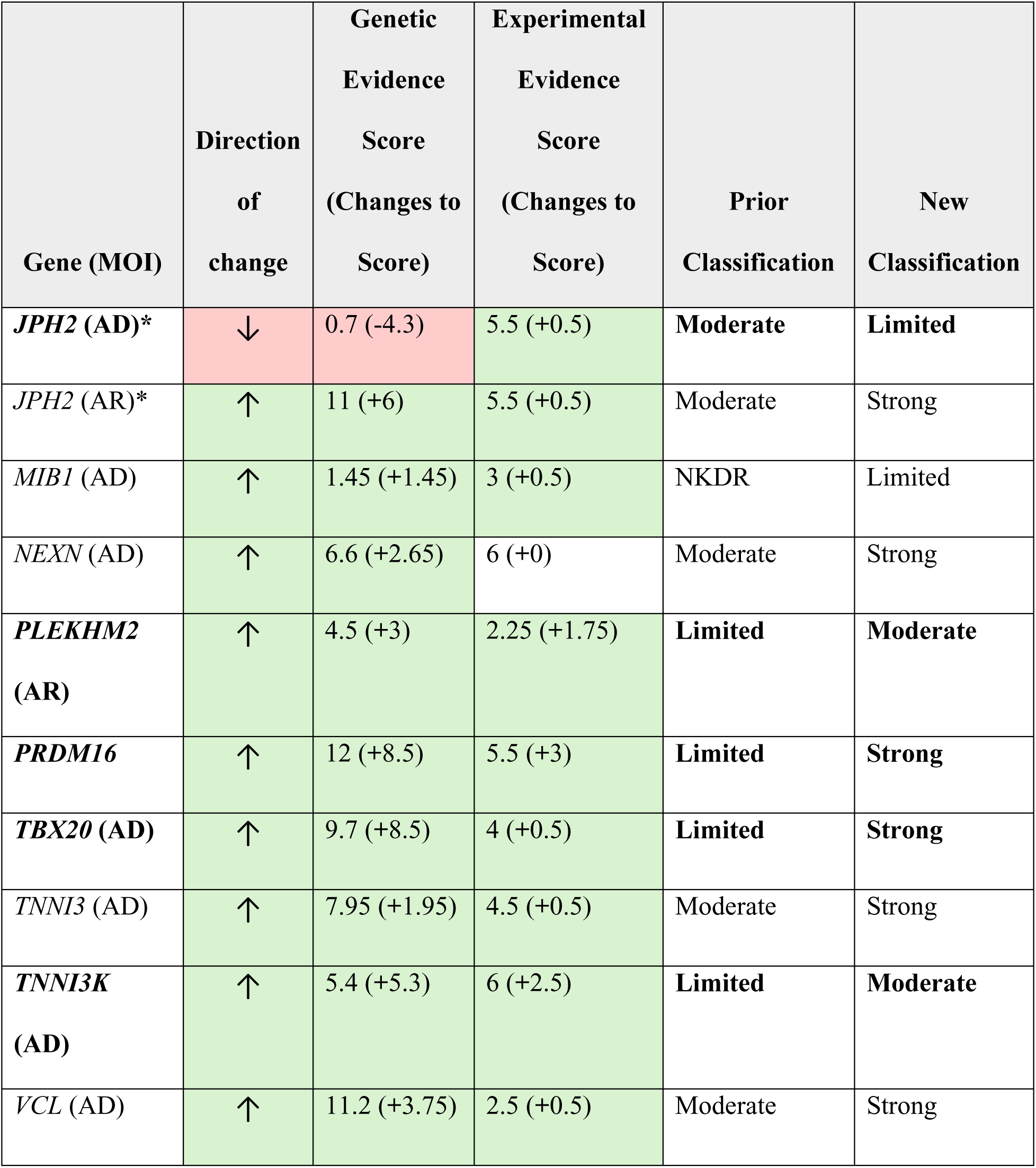
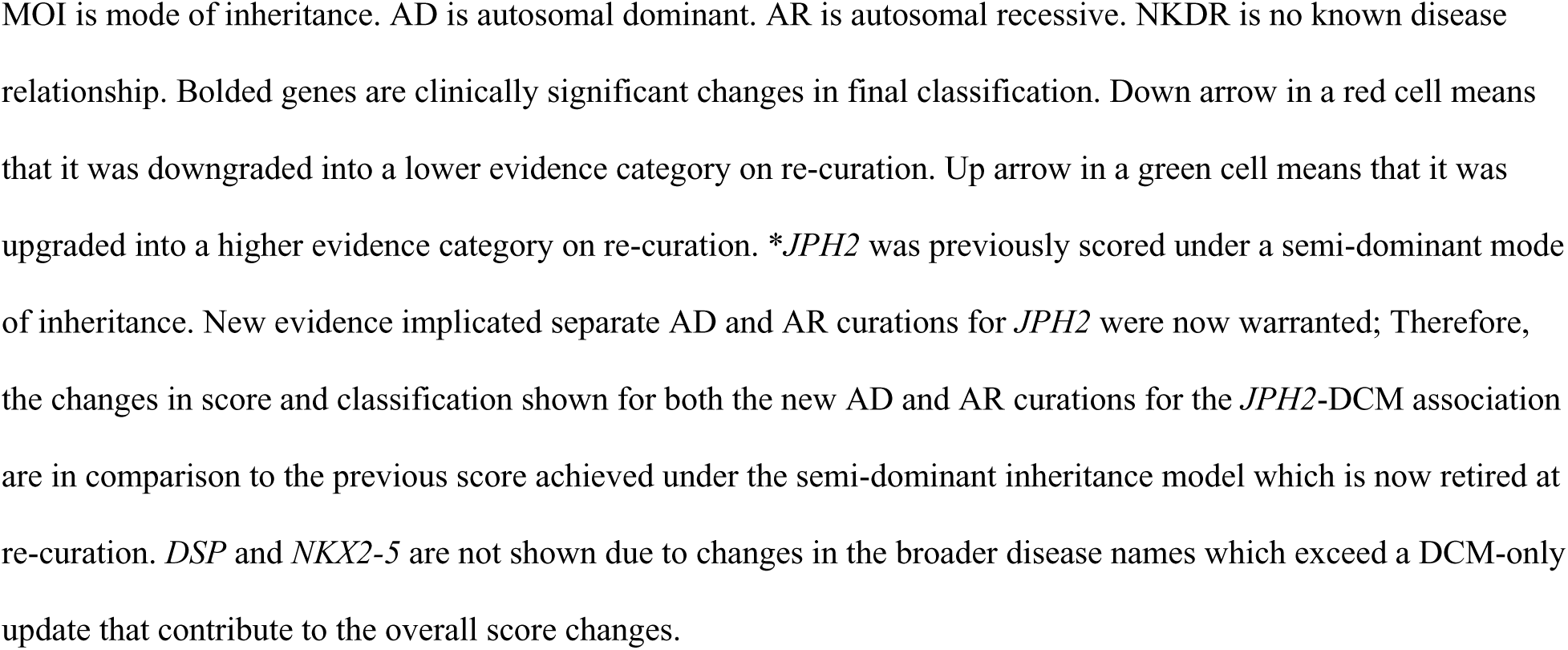
Score changes of re-curations of dilated cardiomyopathy gene-MOI curations that resulted in a change in classification upon updated evidence review.

| Gene (MOI) | Direction of change | Genetic Evidence Score (Changes to Score) | Experimental Evidence Score (Changes to Score) | Prior Classification | New Classification |
| --- | --- | --- | --- | --- | --- |
| *JPH2* (AD)* | ↓ | 0.7 (-4.3) | 5.5 (+0.5) | Moderate | Limited |
| *JPH2* (AR)* | ↑ | 11 (+6) | 5.5 (+0.5) | Moderate | Strong |
| *MIB1* (AD) | ↑ | 1.45 (+1.45) | 3 (+0.5) | NKDR | Limited |
| *NEXN* (AD) | ↑ | 6.6 (+2.65) | 6 (+0) | Moderate | Strong |
| *PLEKHM2* (AR) | ↑ | 4.5 (+3) | 2.25 (+1.75) | Limited | Moderate |
| *PRDM16* | ↑ | 12 (+8.5) | 5.5 (+3) | Limited | Strong |
| *TBX20* (AD) | ↑ | 9.7 (+8.5) | 4 (+0.5) | Limited | Strong |
| *TNNI3* (AD) | ↑ | 7.95 (+1.95) | 4.5 (+0.5) | Moderate | Strong |
| *TNNI3K* (AD) | ↑ | 5.4 (+5.3) | 6 (+2.5) | Limited | Moderate |
| *VCL* (AD) | ↑ | 11.2 (+3.75) | 2.5 (+0.5) | Moderate | Strong |

MOI is mode of inheritance. AD is autosomal dominant. AR is autosomal recessive. NKDR is no known disease relationship. Bolded genes are clinically significant changes in final classification. Down arrow in a red cell means that it was downgraded into a lower evidence category on re-curation. Up arrow in a green cell means that it was upgraded into a higher evidence category on re-curation. \**JPH2* was previously scored under a semi-dominant mode of inheritance. New evidence implicated separate AD and AR curations for *JPH2* were now warranted; Therefore, the changes in score and classification shown for both the new AD and AR curations for the *JPH2*-DCM association are in comparison to the previous score achieved under the semi-dominant inheritance model which is now retired at re-curation. *DSP* and *NKX2-5* are not shown due to changes in the broader disease names which exceed a DCM-only update that contribute to the overall score changes.

### Definitive, Strong, and Moderate Classifications

Across the 35 high evidence curations, 23 were under an AD and 12 under an AR MOI (Figure 1; Figure 3). Twenty-six total high evidence genes were classified as Definitive (n=16) or Strong (n=10), seven of which were newly evaluated AR curations (*JPH2*, *LDB3*, *LMOD2, PPP1R13L, NRAP*, *PPA2, TNNI3*; Figure 3). Nine curations were classified as Moderate evidence for DCM, including four newly evaluated AR curations (*BAG5*, *FLII*, *MYZAP*, *RPL3L*), one new AD curation (*MYLK3)*, and one re-appraised AR curation (*PLEKHM2*). Upon re-appraisal, all genes previously classified as Definitive maintained that classification and narrative summaries were refined and published online (www.clinicalgenome.org). Four genes (five unique curations) previously classified as Moderate achieved a Strong classification at updated assessment (*JPH2*-AR, *NEXN, TNNI3-*AR, *TNNI3*-AD*, VCL*). Four previously Limited evidence genes increased to high evidence categories at re-appraisal, including two now rated as Moderate (*TNNI3K*, *PLEKHM2*) and two as Strong (*PRDM16, TBX20*; Table 2).

**Figure 3.**
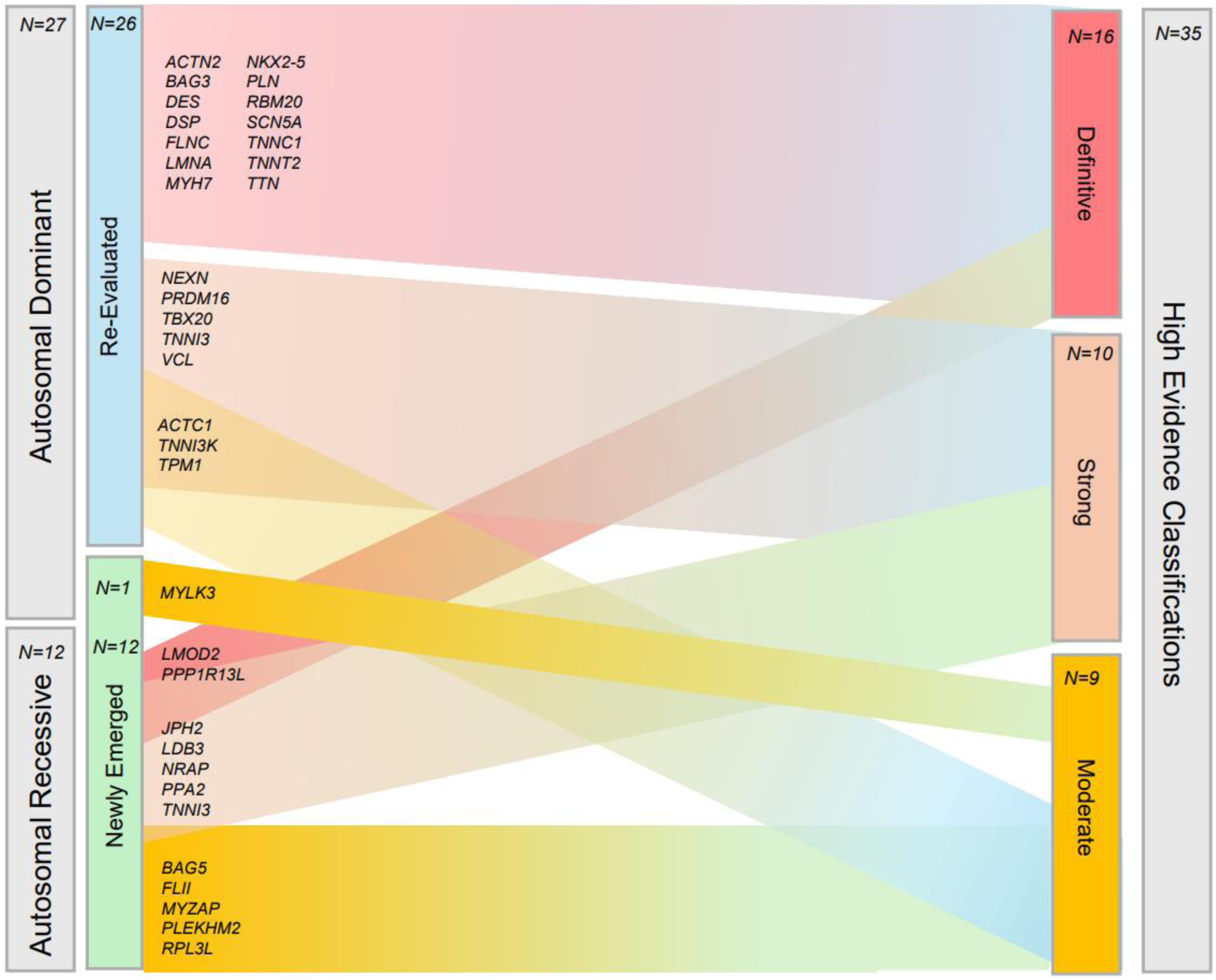
Expanding modes of inheritance of the emerging genetic architecture of high evidence genes in dilated cardiomyopathy. A total of 35 genes that were newly evaluated or re-evaluated under both autosomal dominant and autosomal recessive modes of inheritance achieved a high evidence classification of Moderate, Strong, or Definitive. The colored bars connecting re-evaluated genes (blue box) and newly emerged genes (green box) on the left are connected to the final classification of Definitive (red), Strong (orange), or Moderate (yellow) on the right. The specific gene names in each of these categories are listed on the left hand side in their respective sections, grouped by inheritance mode and whether it is newly evaluated or re-evaluated in the reported curation cycle.

Although eight of the genes classified as Strong have had independent reports over a 3-year period to meet the minimum criteria for Definitive, the panel assigned those curations as Strong to allow additional evidence to accumulate before upgrading, consistent with the DCM GCEP’s conservative application of the framework. This included genes with newly split dual MOI curations (*JPH2*-AR, *LDB3*-AR, *TNNI3*-AD, *TNNI3*-AR) and those with relationships with additional cardiovascular phenotypes where experimental evidence has not demonstrated mechanistic clarity for associated phenotypes (*NEXN, PRDM16, TBX20, VCL*).

### Limited Evidence Classifications

The Limited evidence category was again the largest with 29 total curations: 5 under an AR MOI and 24 AD. This evidence category is also the most heterogenous in overall amount and distribution of evidence points given, ranging from a low total evidence score of 0.6 (*GATA6*) to a high of 10.15 (*MYBPC3*-AD) (Figure 2). More than a third (n=10) of Limited evidence genes accrued < 0.5 human genetic evidence points, with seven genes having as low as 0-0.1 points from published clinical data (Figure 2). These low scoring Limited evidence genes either had unchanged or reduced points after re-evaluation, as in the recuration of *CTF1, DTNA, NEBL, PSEN2,* and *SGCD*.

Seven new curations were scored as Limited for DCM, including six new genes (*CDH2-*AD*, FBXO32-*AR*, GET3-*AR*, RYR2-*AD*, TMOD1-*AR). The seventh gene, *MYBPC3,* evaluated only as AD in the first appraisal,^3^ was also evaluated in this update for an AR MOI. Both AR and AD MOIs for *MYBPC3* were classified as Limited even though the clinical genetic and experimental evidence for *MYBPC3-*AD achieved a cumulative score within the quantitative range for a Moderate classification at the prior and current appraisals (10.05 points total; Table 1, Figure 2). The consensus of the GCEP was again to downgrade to Limited evidence classification for a monogenic cause of DCM due to multiple singleton and/or non-segregating missense variants accumulating points without substantial DCM-specific experimental evidence to augment these reports to a high evidence relationship.

Since the prior assessment, new evidence also supported a curation of unique AD and AR MOIs for *JPH2* to replace the previous semi-dominant assessment, with new clinical genetic evidence supporting an AR MOI for *JPH2*-DCM, prompting the separation of these unique MOI mechanisms for this gene. A semi-dominant MOI framework applies when a phenotype occurs with increasing severity under a spectrum of mono- and bi-allelic presentation rather than AD and AR having independent mechanisms. When considering new evidence, curating an AR MOI for *JPH2* yielded a Strong classification and the separate curation of an AD MOI curation for *JPH2* resulted in a Limited classification (Table 2), a downgrade from the previous Moderate assignment.

### No Known Disease Relationship (NKDR) and Disputed Classifications

With no published human cases meeting the phenotypic criteria and/or harboring plausible variants of clinical relevance, four curations were noted as NKDR for DCM including two recurated (*LRRC10, NPPA*) and two newly evaluated genes (*RTKN2, VEZF1;* Table 1). All four of the previously “Disputed” genes, that is those that lack genetic and/or experimental evidence to suggest a monogenic relationship with DCM, had scores reviewed and updated as applicable, but remained disputed upon reassessment with current evidence (*MYL3*, *PDLIM3, PKP2, PSEN1*; Table 1). We note that a new curation for *MYL3* under an AR MOI was considered; however with only one published consanguineous pedigree^21^ and with an established relationship with HCM,^7^ this was deemed insufficient to warrant an independent assessment at this time. Should new evidence emerge, a separate curation of *MYL3* for DCM under an AR MOI may be appropriate.

### Classification Changes of Previously Evaluated Genes

For the 51 re-appraised genes, 42 did not change classification. The remaining nine genes resulted in ten unique curations with changed clinical validity classifications (Table 2). Of these, four resulted in clinically significant upgrades (*PLEKHM2* and *TNNI3K* from Limited to Moderate; *TBX20* and *PRDM16* from Limited to Strong). Under a new AD MOI, *JPH2*-AD was downgraded from Moderate to Limited due to a reduction of 4.3 points in clinical genetic evidence for a genetic evidence score of 0.7. This downgrade was paired with the separately assessed *JPH2*-AR curation noted above to result in a Strong classification. With substantial amounts of new evidence since the prior assessment, the largest changes in overall quantitative score and final classifications occurred with *TBX20*^22, 23^ and *PRDM16*.^24–28^

The *DSP* and *NKX2-5* classifications were updated to Definitive but are not shown in Table 2 as they were, in part, from classifications beyond that of only DCM, as primarily curated by other ClinGen GCEPs. DCM clinical genetic and experimental evidence points contributing significantly to the final scores within disease relationships of *DSP*-arrhythmogenic cardiomyopathy with wooly hair and keratoderma (MONDO 0011581) and *NKX2.5*-related congenital, conduction, and myopathic heart disease (MONDO 0800441), as described in the ClinGen narrative summaries (www.clinicalgenome.org).

## DISCUSSION

An updated, evidence-based assessment of clinically relevant genes in DCM was conducted based on a recent remarkable expansion of published literature. In the five years since the curation effort in 2019-2020, an increase of 139 candidate genes for a total of 406 in the initial list were identified using the same database query methodology from the prior work.^3^ We report clinically relevant updates, most notably an increase from 19 to 35 high evidence curations with over one-third novel for an autosomal recessive (AR) mode of inheritance.

The most significant change was a five-fold increase in curations under an AR inheritance model compared to the initial curation, to a total of 20. Many of the new AR curations were based upon analyses from large cohorts of pediatric-onset cardiomyopathy, many observed in consanguineous families,^29–31^ revealing novel insights. In the curation of AR inheritance, the GCEP utilized a conservative approach, requiring parental data to maximize the confidence that an individual carrying only one (heterozygous) rare variant had no evidence of a DCM phenotype. In the absence of such data, points were reduced. The much greater availability of published trio and family data permitted these genes to be considered. In the case of related, consanguineous families, variant homozygosity was frequently observed, that is, where both maternally and paternally inherited alleles contributed the same rare variant to the offspring.

Statistically these homozygous observations of very rare, and in some cases novel, variants would have been extremely unlikely to have been observed by chance except in consanguineous pedigrees; therefore, as another means to avoid over-scoring evidence, points were reduced if consanguinity was reported or not explicitly stated to be ruled out in homozygous observations.

Consideration of both AD and AR MOIs for *JPH2, LDB3, MYBPC3,* and *TNNI3* revealed potential mechanistic differences (Supplemental Table 2), underscoring the impact and robustness of the ClinGen curation framework^15^ in illuminating important variant characteristics. This was evident in the assessment of *TNNI3,* previously only curated for an AD MOI. Since the prior curation, the *TNNI3-*AD curation added 2 points evidence to the total score, resulting in an upgraded classification from Moderate to Strong.^32, 33^ Most of the scored variants for *TNNI3-*AD were non-truncating, which accrued lower points per variant than predicted null variants based upon the default scores set in the current SOP. Multiple variant observations were therefore required to accomplish a high total cumulative score, and in the case of *TNNI3-*AD, 22 non-truncating variants were evaluated which cumulatively contributed 6.95 points to the total genetic evidence score. In contrast, the new AR MOI curation of *TNNI3* accumulated the maximum score of 12 genetic evidence points in addition to 4.5 experimental evidence points, reaching Strong evidence with nearly all the variants having a predicted null consequence.

Multiple observations of the c.204del (p.Arg69Alafs*8)^32, 34–36^ and c.292C>T (p.Arg98*)^33, 34^ rare variants were reported and scored in pediatric DCM, in addition to several others.

The clinical validity assessment of a gene-disease relationship precedes the application of clinical variant classifications standards defined by ACMG/AMP, which elevates the clinical relevance of “high evidence” assignments.^19^ The work herein defines a gene set for which current standards of variant classifications can be applied in clinical practice. Notably *JPH2*, classified as Moderate evidence in the prior publication under a semi-dominant MOI, has been upgraded with a separate AR curation rated as Strong but downgraded for an AD MOI to Limited, demonstrating the plasticity and complexity of the evolving DCM genetics knowledgebase. The Limited clinical genetic evidence (0.7 points) for *JPH2-*AD in DCM raises the question of whether heterozygous *JPH2* variants represent only carrier status or if an intermediate or monoallelic effect may also be at play. The expert panel noted the downgraded classification for *JPH2*-AD will affect clinical classifications and thus may trigger cause for updated clinical correlation.

This reassessment extended the prior curation effort to reveal a markedly expanded and diverse spectrum of disease ontologies across implicated genes, underscoring the substantial genetic complexity of DCM (Figure 1),^3^ as shown by this work and reviewed by others.^29^ The previous curation had 10 gene ontologies represented across high evidence DCM genes; this has now been expanded with eight additional gene ontologies, including actin-binding, transcription factors, lysosomal function and autophagy, P53 inhibition, ribosomal assembly, contractility regulation and mitochondrial energy production. With now multiple MOIs also emerging for DCM causation (Figure 3), the robustness of the ClinGen process revealed patterns within gene ontology groups, for example with all actin-binding genes operating under an -AR MOI for DCM (Figure 1). We re-emphasize that the Limited evidence classification again had the most genes, an expanded list now numbering 29. The ClinGen gene curation framework has been designed to rigorously evaluate monogenic cause for the utility of clinical diagnostics and familial risk stratification, where the presence or absence of a variant(s) in a single gene informs the diagnosis or risk, as applicable, of genetic DCM. While useful and parsimonious, a monogenic approach is likely incomplete for some DCM probands, and perhaps substantially so for selected genes or for some pedigrees that may be better analyzed inclusive of polygenic risk, intermediate effect variants,^37^ or an oligogenic model.^38^ Whether genes classified as Limited evidence may fit better within an oligogenic or more complex genetic model remains to be established.

An example was the curation of *MYPBC3* for DCM, a gene primarily associated with HCM,^5–7^ which in the 2019-2020 curation accumulated substantial variant-level points from rare missense variants in numerous singleton cases. This increased the DCM-specific point-based score for the AD MOI for *MYBPC3* into the Moderate range. The *MYPBC3* point-based score for DCM was increased even more in the 2025 curation when applying the same standards. For the 2025 assessment, published cases had also accumulated with biallelic *MYBPC3* variants so an AR MOI was evaluated. However, due to the lack of evidence of variants segregating in DCM pedigrees to validate a monogenic disease mechanism, and no statistical enrichment in DCM populations,^39^ the classifications assigned were maintained as Limited for both *MYBPC3*-AD and *MYBPC3*-AR curations. For *MYBPC3*-AD, this required the GCEP to override the quantitative score, as the consensus of the expert panel was that while these variants may contribute to a DCM phenotype, the evidence was insufficient to support a monogenic cause of DCM. Whether *MYBPC3* or other similarly scored genes should be considered as DCM risk-associated genes remains an open question. Measures of non-monogenic risk have yet to be defined for clinical translation.

## CONCLUSION

The ClinGen DCM GCEP conducted an updated gene curation, further expanding our understanding of the complex and evolving genetic architecture of DCM. We have reported several new genes, updated previous genes, and provided numerous new curations under an AR MOI. Continuous rigorous curation of monogenic cause will be essential as DCM genetic literature evolves, and expanded frameworks that incorporate mechanisms beyond monogenic inheritance may help to further clarify the complex genetic architecture of DCM.

## Data Availability

All of the data are available on the ClinGen website.

https://clinicalgenome.org/

## ACKNOWLEDGEMENTS

The authors thank Stephanie Schulte, MLIS, Associate Professor and Head of Research and Education Services at the Health Sciences Library at The Ohio State University, for her assistance in developing a systematic and comprehensive approach to developing the initial gene list from the OMIM, Gene, and GenBank databases.

## NON-STANDARD ABBREVIATIONS AND ACRONYMS

ACMG: American College of Medical Genetics
AD: Autosomal dominant
AMP: Association of Molecular Pathology
AR: Autosomal recessive
ARVC: Arrhythmogenic right ventricular cardiomyopathy
ClinGen: Clinical Genome Resource
DCM: Dilated cardiomyopathy
GCEP: Gene curation expert panel
HCM: hypertrophic cardiomyopathy
LP: Likely pathogenic
MOI: Mode of inheritance
NKDR: No known disease relationship
P: Pathogenic
SOP: Standard Operating Procedures
VUS: variant of uncertain significance

